# Discordant SARS-CoV-2 PCR and Rapid Antigen Test Results When Infectious: A December 2021 Occupational Case Series

**DOI:** 10.1101/2022.01.04.22268770

**Authors:** Blythe Adamson, Robby Sikka, Anne L. Wyllie, Prem Premsrirut

## Abstract

The performance of Covid-19 diagnostic tests must continue to be reassessed with new variants of concern. The objective of this study was to describe the discordance in saliva SARS-CoV-2 PCR and nasal rapid antigen test results during the early infectious period. We identified a high-risk occupational case cohort of 30 individuals with daily testing during an Omicron outbreak in December 2021. Based on viral load and transmissions confirmed through epidemiological investigation, most Omicron cases were infectious for several days before being detectable by rapid antigen tests.

---

Routine workplace Covid-19 surveillance testing has been key to reopening in-person businesses with job functions that have high-risk for SARS-CoV-2 transmission. Yet, the real world performance of Covid-19 diagnostic tests needs to be reassessed for each new variant of concern. The FDA recently updated guidance stating that antigen tests may be less sensitive for the detection of Omicron than previous SARS-CoV-2 variants.^1^ However, laboratory experiments cannot fully replace clinical study evaluations using patient samples through the course of SARS-CoV-2 infection. Omicron has been shown to infect faster and more efficiently than Delta in human bronchus, but with less severe infection in lung,^2^ translating to symptom increase of sore throats and decrease of loss of taste and smell, better detected by saliva than nasal swabs.^3-5^ To date, the viral dynamics and test performance in the Omicron early infection period have not been described in detail, as it requires cohorts receiving near daily testing to identify cases prior to symptom onset.

## METHODS

This retrospective cohort study identified individuals in occupational safety programs who were diagnosed with Covid-19 between December 1-31, 2021 during Omicron outbreaks at five workplaces in New York, NY, Los Angeles, CA, and San Francisco, CA. The populations were fully vaccinated by employer mandate and highly boosted by choice. To isolate the window of acute infection, cases were included if they were receiving daily testing at the time of diagnosis, had paired SARS-CoV-2 quantitative reverse-transcriptase polymerase chain reaction (RT-qPCR) test results and rapid antigen test results on Day 0 or 1 relative to first positive specimen collection date and were excluded if missing a recent negative test. The primary outcome of interest was discordance between saliva PCR and nasal rapid antigen test results during the early period with viral load levels corresponding to infectious risk of transmission. A Kaplan Meier analysis was performed to estimate the median time from first PCR positive to first rapid antigen positive test result.

## RESULTS

Among hundreds of workplace-detected Covid-19 cases in December 2021, we identified 30 individuals with 62 matched pairs of rapid antigen and positive PCR results from specimens collected at the same time. The S-gene dropout associated with Omicron was observed in 29 of 30 cases. Viral dynamics and discordance in test results are shown in Figure 1. Four cases were confirmed to have transmitted the virus between false-negative antigen tests, with saliva PCR cycle threshold (Ct) values between 23-28 for the N gene. On Days 0 and 1, all rapid antigen tests produced false-negative results, despite 28 of 30 pairs having infectious viral load within the range of confirmed Omicron transmissions in the cohort (Ct < 29). The median time from first positive PCR to first detectable antigen positive was 3 days (95% Confidence Interval: 2-NA). After infection was detected, a subgroup (n=5) who received daily saliva PCR, nasal swab PCR, and nasal swab rapid antigen testing showed viral load peaked in saliva 1-2 days before nasal tests (Supplemental Table 1). All individuals in the cohort developed symptoms within two days of the first PCR positive test.

**Figure 1.**
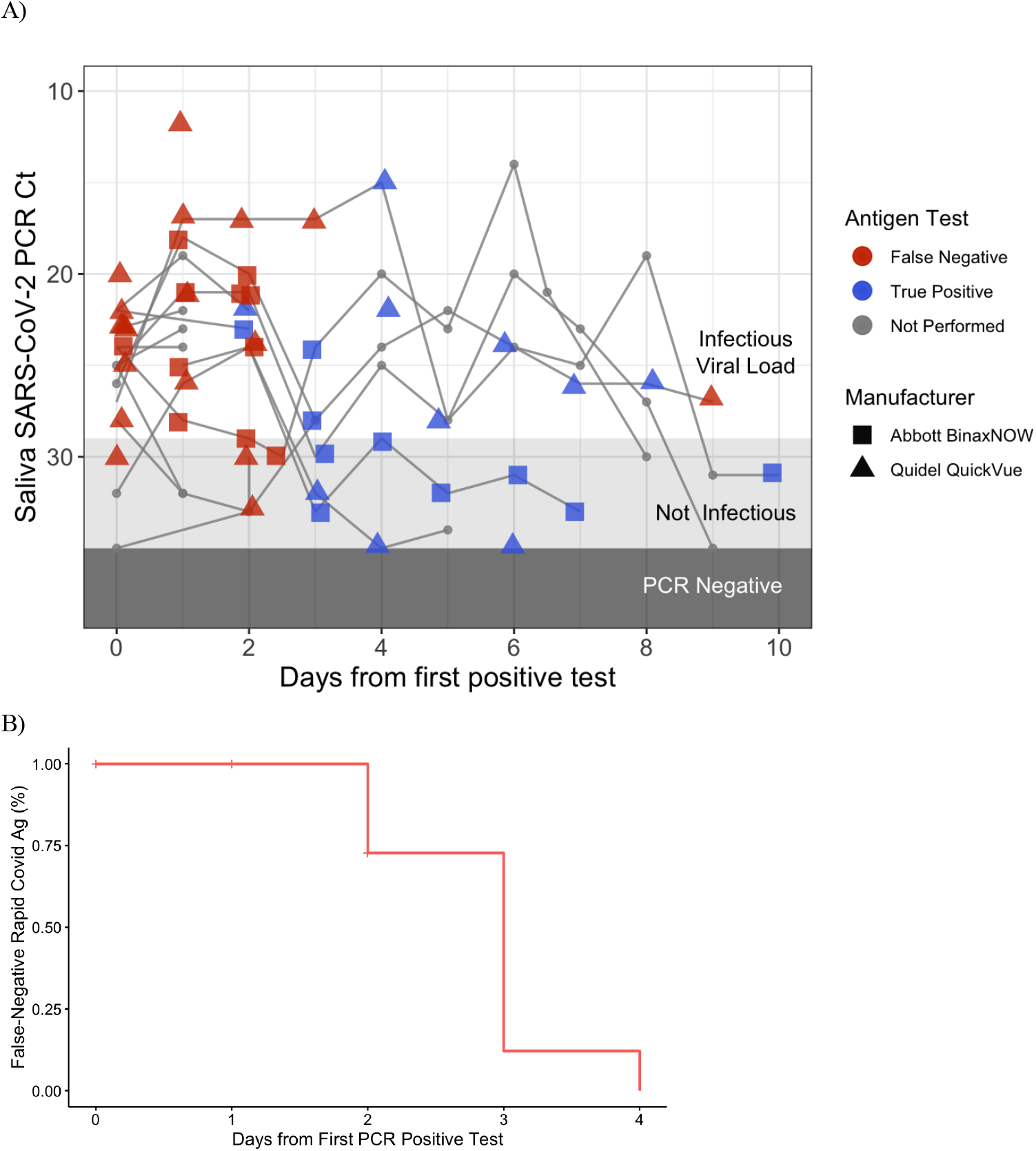
Discordance in PCR and Rapid Antigen Test Results. Shown are Cycle Threshold (Ct) counts and rapid antigen test results from paired samples of infected patients (Panel A). Also shown is the Kaplan Meier analysis of time from positive PCR to positive rapid antigen test (Panel B).

## DISCUSSION

We found that rapid antigen tests lagged in the ability to detect Covid-19 during an early period of disease when most individuals were infectious with Omicron and four transmissions were confirmed. The policy implication is that rapid antigen tests may not be as fit-for-purpose in routine workplace screening to prevent asymptomatic spread of Omicron, compared to prior variants,^6^ given the shorter time from exposure to infectiousness and lower infectious doses sufficient for transmission. These findings are consistent with population-level Omicron epidemiology studies showing shorter serial intervals between cases and faster rates of community spread. Despite the small numbers of individuals included in this study, the findings are uniquely valuable because of the early detection of Omicron infection in frequent workplace Covid-19 testing to prevent spread. In real-world antigen testing, the limit of detection was substantially lower than manufacturers have reported to the FDA based on laboratory validation.

## Data Availability

All de-identified data and the code for analysis is available on GitHub at https://github.com/blythejane/covid_safety.

## Acknowledgements

We thank the COVID-19 Sports and Society Working Group, workplace COVID Safety Managers, Harlan Krumholz, MD and Nathan Grubaugh, PhD from Yale University, and the operational team, including Charlene Speyer and Lisa Kussell from Infectious Economics.

## Supplementary Appendix

### Study Design

The data reported here represent a convenience sample of employees at a selection of workplaces that partner with Infectious Economics LLC for Covid-19 prevention, including corporate offices, entertainment, retail trade, and manufacturing. The study period ran December 1, 2021 to December 31, 2021. Clinical samples were obtained by self-collection observed by a trained COVID Safety Manager. Employer-provided testing was preventatively provided to enable in-person workplace safety during times of high community prevalence and/or recent COVID exposures in the workplace. Because the sensitivity of tests may vary during the course of an infection, we evaluated concordance of PCR and rapid antigen test results in matched samples over time.

### Study Oversight

In accordance with the guidelines, this work with de-identified samples was approved for research not involving human subjects by the SUNY Downstate Institutional Review Board & Privacy Board (1603504-6) under the title “Pilot program for instituting massive COVID19 surveillance screening in schools and the workplace.”

### Covid-19 Testing and Sequencing

The rapid antigen tests kits used were Quidel QuickVue At-Home OTC COVID-19 Test and Abbott BinaxNOW COVID-19 Antigen Self-Test. PCR to detect SARS-CoV-2 RNA was the ThermoFisher Combo Kit, allowing for the ability to detect an S-gene dropout. The time from specimen collection to results returned was <30 minutes for antigen tests and ranged 6-12 hours for PCR. A subset of specimens received whole genome sequencing in a broader epidemiology investigation of clusters to further improve workplace safety policies. Samples were classified as Omicron or Delta based on whole genome sequencing data, diagnostic PCR target failures, and sampling dates. In whole genome sequencing, RNA was extracted and confirmed as SARS-CoV-2 positive by RT-qPCR with the Thermo Fisher TaqPath SARS-CoV-2 assay. Next Generation Sequencing with the Illumina COVIDSeq ARTIC primer set2 was used for viral amplification.

### Outcomes

The primary outcome of interest was concordance between PCR and rapid antigen test results, dependent on time and Ct values corresponding to infectiousness. We defined the index as the specimen collection date for the first PCR test with detectable SARS-CoV-2 with Ct <35, the event being the first positive rapid antigen test result, and censoring at the most recent antigen test date. Based on observed transmission events in these workplaces that were confirmed by contact tracing and genomic epidemiology investigation, we defined infectious viral load as corresponding to Ct values <29 in this analysis.

### Statistical analysis

A Kaplan Meier analysis was performed to estimate the median time from PCR positive to rapid antigen positive and time-dependent probability of a false negative rapid antigen test. We defined the index as the specimen collection date for the first PCR test with detectable SARS-CoV-2 with Ct <35, the event being the first positive rapid antigen test result, and censoring at the most recent antigen test date. Analyses were conducted in R 4.1.2.

**Supplementary Table 1.**
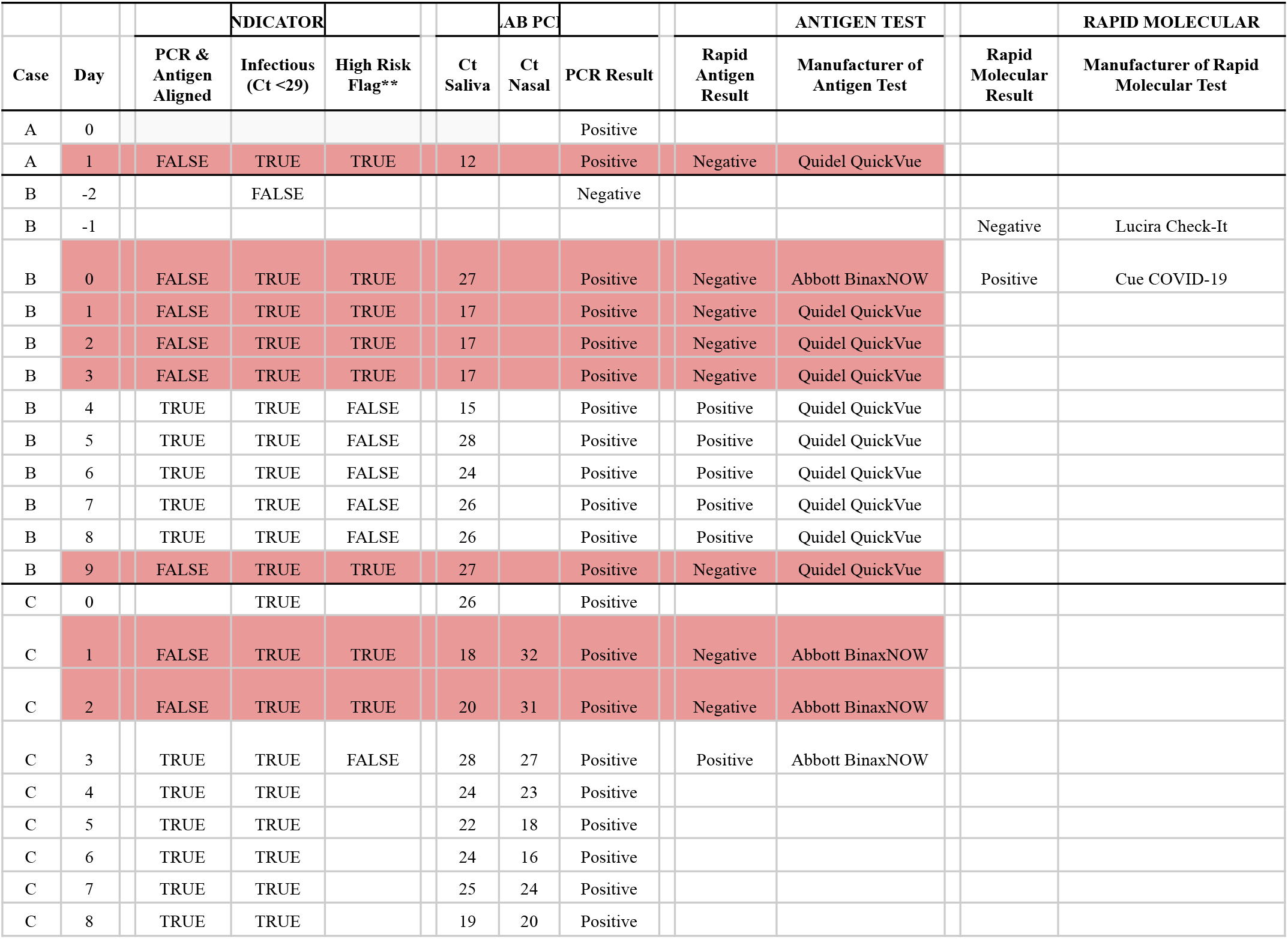
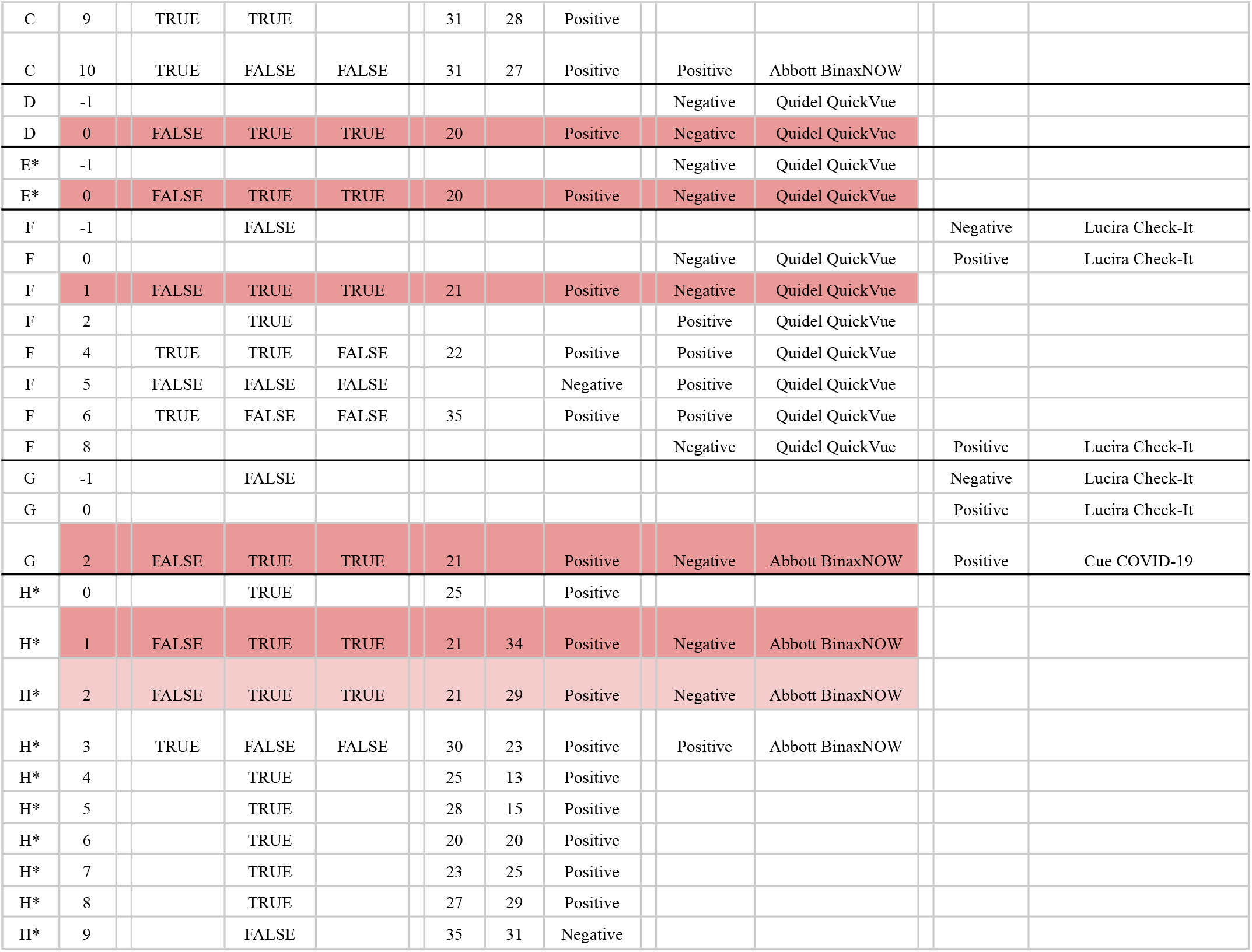
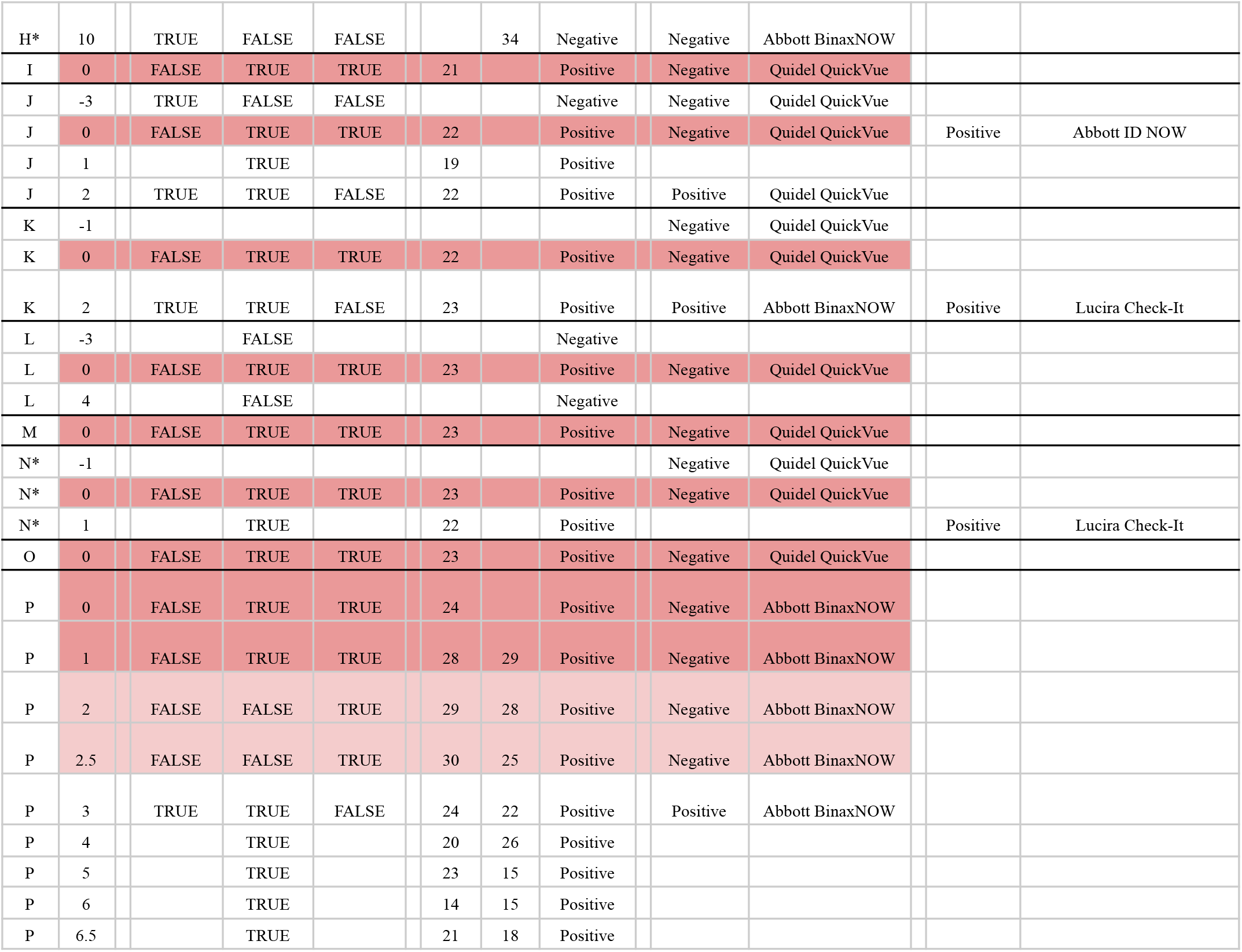
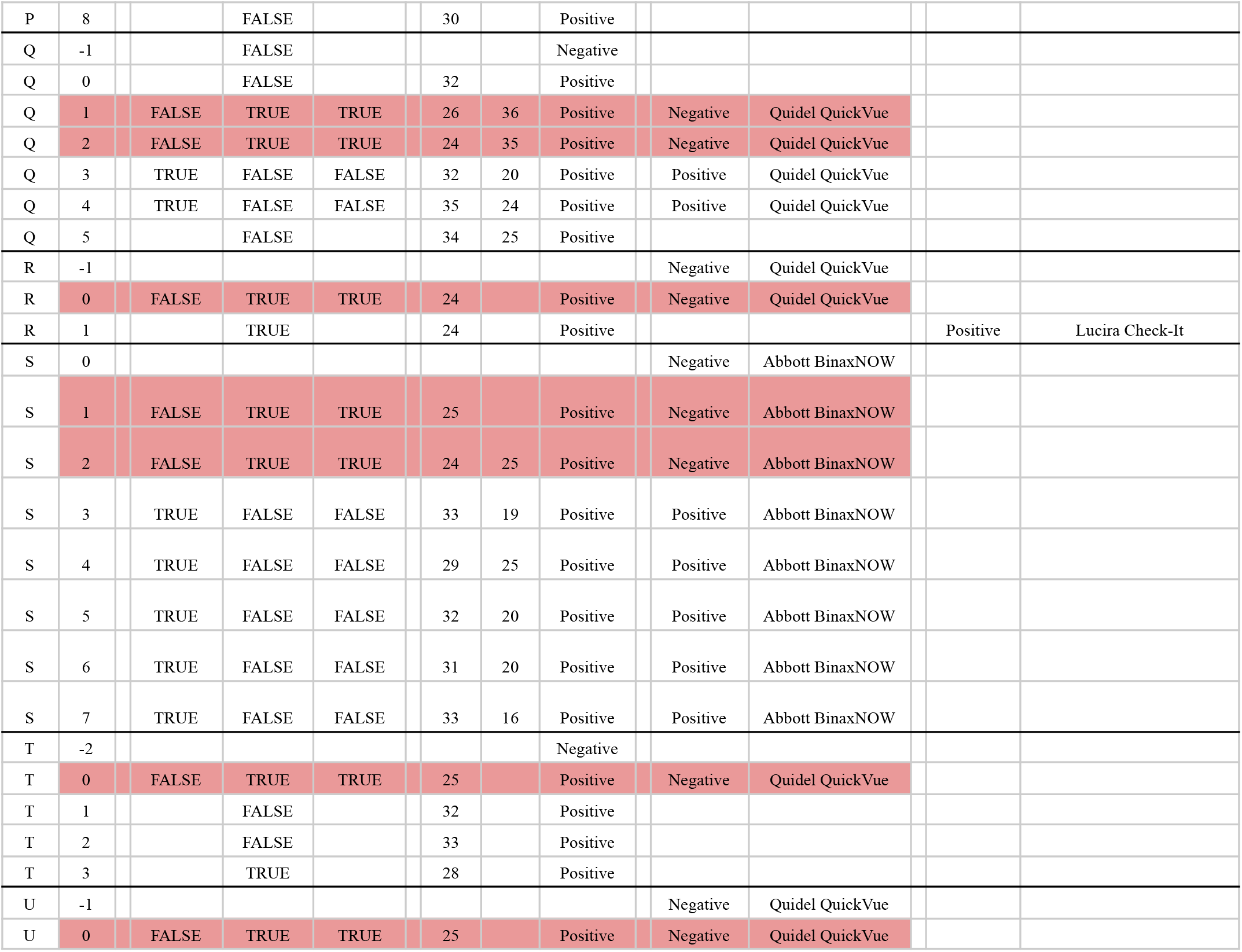
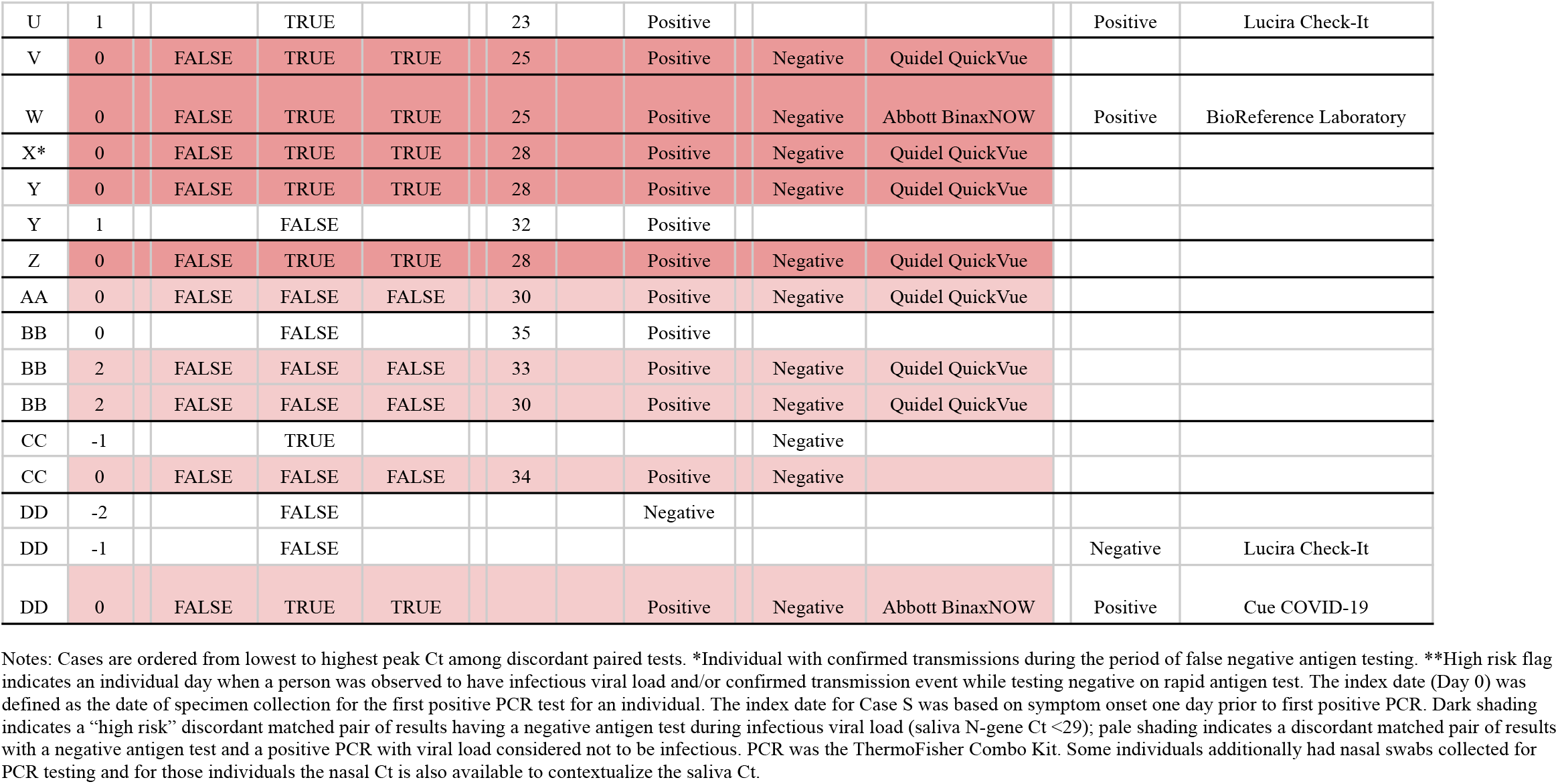
Covid-19 cases with discordant test results from paired PCR and antigen tests in workplace safety programs

|  |  |  | INDICATOR |  |  | LAB PC |  |  | ANTIGEN TEST |  | RAPID MOLECULAR |
| --- | --- | --- | --- | --- | --- | --- | --- | --- | --- | --- | --- |
| Case | Day | PCR & Antigen Aligned | Infectious (Ct <29) | High Risk Flag** | Ct Saliva | Ct Nasal | PCR Result | Rapid Antigen Result | Manufacturer of Antigen Test | Rapid Molecular Result | Manufacturer of Rapid Molecular Test |
| A | 0 |  |  |  |  |  | Positive |  |  |  |  |
| A | 1 | FALSE | TRUE | TRUE | 12 |  | Positive | Negative | Quidel QuickVue |  |  |
| B | -2 |  | FALSE |  |  |  | Negative |  |  |  |  |
| B | -1 |  |  |  |  |  |  |  |  | Negative | Lucira Check-It |
| B | 0 | FALSE | TRUE | TRUE | 27 |  | Positive | Negative | Abbott BinaxNOW | Positive | Cue COVID-19 |
| B | 1 | FALSE | TRUE | TRUE | 17 |  | Positive | Negative | Quidel QuickVue |  |  |
| B | 2 | FALSE | TRUE | TRUE | 17 |  | Positive | Negative | Quidel QuickVue |  |  |
| B | 3 | FALSE | TRUE | TRUE | 17 |  | Positive | Negative | Quidel QuickVue |  |  |
| B | 4 | TRUE | TRUE | FALSE | 15 |  | Positive | Positive | Quidel QuickVue |  |  |
| B | 5 | TRUE | TRUE | FALSE | 28 |  | Positive | Positive | Quidel QuickVue |  |  |
| B | 6 | TRUE | TRUE | FALSE | 24 |  | Positive | Positive | Quidel QuickVue |  |  |
| B | 7 | TRUE | TRUE | FALSE | 26 |  | Positive | Positive | Quidel QuickVue |  |  |
| B | 8 | TRUE | TRUE | FALSE | 26 |  | Positive | Positive | Quidel QuickVue |  |  |
| B | 9 | FALSE | TRUE | TRUE | 27 |  | Positive | Negative | Quidel QuickVue |  |  |
| C | 0 |  | TRUE |  | 26 |  | Positive |  |  |  |  |
| C | 1 | FALSE | TRUE | TRUE | 18 | 32 | Positive | Negative | Abbott BinaxNOW |  |  |
| C | 2 | FALSE | TRUE | TRUE | 20 | 31 | Positive | Negative | Abbott BinaxNOW |  |  |
| C | 3 | TRUE | TRUE | FALSE | 28 | 27 | Positive | Positive | Abbott BinaxNOW |  |  |
| C | 4 | TRUE | TRUE |  | 24 | 23 | Positive |  |  |  |  |
| C | 5 | TRUE | TRUE |  | 22 | 18 | Positive |  |  |  |  |
| C | 6 | TRUE | TRUE |  | 24 | 16 | Positive |  |  |  |  |
| C | 7 | TRUE | TRUE |  | 25 | 24 | Positive |  |  |  |  |
| C | 8 | TRUE | TRUE |  | 19 | 20 | Positive |  |  |  |  |
| C | 9 | TRUE | TRUE |  | 31 | 28 | Positive |  |  |  |  |
| C | 10 | TRUE | FALSE | FALSE | 31 | 27 | Positive | Positive | Abbott BinaxNOW |  |  |
| D | -1 |  |  |  |  |  |  | Negative | Quidel QuickVue |  |  |
| D | 0 | FALSE | TRUE | TRUE | 20 |  | Positive | Negative | Quidel QuickVue |  |  |
| E* | -1 |  |  |  |  |  |  | Negative | Quidel QuickVue |  |  |
| E* | 0 | FALSE | TRUE | TRUE | 20 |  | Positive | Negative | Quidel QuickVue |  |  |
| F | -1 |  | FALSE |  |  |  |  |  |  | Negative | Lucira Check-It |
| F | 0 |  |  |  |  |  |  | Negative | Quidel QuickVue | Positive | Lucira Check-It |
| F | 1 | FALSE | TRUE | TRUE | 21 |  | Positive | Negative | Quidel QuickVue |  |  |
| F | 2 |  | TRUE |  |  |  |  | Positive | Quidel QuickVue |  |  |
| F | 4 | TRUE | TRUE | FALSE | 22 |  | Positive | Positive | Quidel QuickVue |  |  |
| F | 5 | FALSE | FALSE | FALSE |  |  | Negative | Positive | Quidel QuickVue |  |  |
| F | 6 | TRUE | FALSE | FALSE | 35 |  | Positive | Positive | Quidel QuickVue |  |  |
| F | 8 |  |  |  |  |  |  | Negative | Quidel QuickVue | Positive | Lucira Check-It |
| G | -1 |  | FALSE |  |  |  |  |  |  | Negative | Lucira Check-It |
| G | 0 |  |  |  |  |  |  |  |  | Positive | Lucira Check-It |
| G | 2 | FALSE | TRUE | TRUE | 21 |  | Positive | Negative | Abbott BinaxNOW | Positive | Cue COVID-19 |
| H* | 0 |  | TRUE |  | 25 |  | Positive |  |  |  |  |
| H* | 1 | FALSE | TRUE | TRUE | 21 | 34 | Positive | Negative | Abbott BinaxNOW |  |  |
| H* | 2 | FALSE | TRUE | TRUE | 21 | 29 | Positive | Negative | Abbott BinaxNOW |  |  |
| H* | 3 | TRUE | FALSE | FALSE | 30 | 23 | Positive | Positive | Abbott BinaxNOW |  |  |
| H* | 4 |  | TRUE |  | 25 | 13 | Positive |  |  |  |  |
| H* | 5 |  | TRUE |  | 28 | 15 | Positive |  |  |  |  |
| H* | 6 |  | TRUE |  | 20 | 20 | Positive |  |  |  |  |
| H* | 7 |  | TRUE |  | 23 | 25 | Positive |  |  |  |  |
| H* | 8 |  | TRUE |  | 27 | 29 | Positive |  |  |  |  |
| H* | 9 |  | FALSE |  | 35 | 31 | Negative |  |  |  |  |
| H* | 10 | TRUE | FALSE | FALSE |  | 34 | Negative | Negative | Abbott BinaxNOW |  |  |
| I | 0 | FALSE | TRUE | TRUE | 21 |  | Positive | Negative | Quidel QuickVue |  |  |
| J | -3 | TRUE | FALSE | FALSE |  |  | Negative | Negative | Quidel QuickVue |  |  |
| J | 0 | FALSE | TRUE | TRUE | 22 |  | Positive | Negative | Quidel QuickVue | Positive | Abbott ID NOW |
| J | 1 |  | TRUE |  | 19 |  | Positive |  |  |  |  |
| J | 2 | TRUE | TRUE | FALSE | 22 |  | Positive | Positive | Quidel QuickVue |  |  |
| K | -1 |  |  |  |  |  |  | Negative | Quidel QuickVue |  |  |
| K | 0 | FALSE | TRUE | TRUE | 22 |  | Positive | Negative | Quidel QuickVue |  |  |
| K | 2 | TRUE | TRUE | FALSE | 23 |  | Positive | Positive | Abbott BinaxNOW | Positive | Lucira Check-It |
| L | -3 |  | FALSE |  |  |  | Negative |  |  |  |  |
| L | 0 | FALSE | TRUE | TRUE | 23 |  | Positive | Negative | Quidel QuickVue |  |  |
| L | 4 |  | FALSE |  |  |  | Negative |  |  |  |  |
| M | 0 | FALSE | TRUE | TRUE | 23 |  | Positive | Negative | Quidel QuickVue |  |  |
| N* | -1 |  |  |  |  |  |  | Negative | Quidel QuickVue |  |  |
| N* | 0 | FALSE | TRUE | TRUE | 23 |  | Positive | Negative | Quidel QuickVue |  |  |
| N* | 1 |  | TRUE |  | 22 |  | Positive |  |  | Positive | Lucira Check-It |
| O | 0 | FALSE | TRUE | TRUE | 23 |  | Positive | Negative | Quidel QuickVue |  |  |
| P | 0 | FALSE | TRUE | TRUE | 24 |  | Positive | Negative | Abbott BinaxNOW |  |  |
| P | 1 | FALSE | TRUE | TRUE | 28 | 29 | Positive | Negative | Abbott BinaxNOW |  |  |
| P | 2 | FALSE | FALSE | TRUE | 29 | 28 | Positive | Negative | Abbott BinaxNOW |  |  |
| P | 2.5 | FALSE | FALSE | TRUE | 30 | 25 | Positive | Negative | Abbott BinaxNOW |  |  |
| P | 3 | TRUE | TRUE | FALSE | 24 | 22 | Positive | Positive | Abbott BinaxNOW |  |  |
| P | 4 |  | TRUE |  | 20 | 26 | Positive |  |  |  |  |
| P | 5 |  | TRUE |  | 23 | 15 | Positive |  |  |  |  |
| P | 6 |  | TRUE |  | 14 | 15 | Positive |  |  |  |  |
| P | 6.5 |  | TRUE |  | 21 | 18 | Positive |  |  |  |  |
| P | 8 |  | FALSE |  | 30 |  | Positive |  |  |  |  |
| Q | -1 |  | FALSE |  |  |  | Negative |  |  |  |  |
| Q | 0 |  | FALSE |  | 32 |  | Positive |  |  |  |  |
| Q | 1 | FALSE | TRUE | TRUE | 26 | 36 | Positive | Negative | Quidel QuickVue |  |  |
| Q | 2 | FALSE | TRUE | TRUE | 24 | 35 | Positive | Negative | Quidel QuickVue |  |  |
| Q | 3 | TRUE | FALSE | FALSE | 32 | 20 | Positive | Positive | Quidel QuickVue |  |  |
| Q | 4 | TRUE | FALSE | FALSE | 35 | 24 | Positive | Positive | Quidel QuickVue |  |  |
| Q | 5 |  | FALSE |  | 34 | 25 | Positive |  |  |  |  |
| R | -1 |  |  |  |  |  |  | Negative | Quidel QuickVue |  |  |
| R | 0 | FALSE | TRUE | TRUE | 24 |  | Positive | Negative | Quidel QuickVue |  |  |
| R | 1 |  | TRUE |  | 24 |  | Positive |  |  | Positive | Lucira Check-It |
| S | 0 |  |  |  |  |  |  | Negative | Abbott BinaxNOW |  |  |
| S | 1 | FALSE | TRUE | TRUE | 25 |  | Positive | Negative | Abbott BinaxNOW |  |  |
| S | 2 | FALSE | TRUE | TRUE | 24 | 25 | Positive | Negative | Abbott BinaxNOW |  |  |
| S | 3 | TRUE | FALSE | FALSE | 33 | 19 | Positive | Positive | Abbott BinaxNOW |  |  |
| S | 4 | TRUE | FALSE | FALSE | 29 | 25 | Positive | Positive | Abbott BinaxNOW |  |  |
| S | 5 | TRUE | FALSE | FALSE | 32 | 20 | Positive | Positive | Abbott BinaxNOW |  |  |
| S | 6 | TRUE | FALSE | FALSE | 31 | 20 | Positive | Positive | Abbott BinaxNOW |  |  |
| S | 7 | TRUE | FALSE | FALSE | 33 | 16 | Positive | Positive | Abbott BinaxNOW |  |  |
| T | -2 |  |  |  |  |  | Negative |  |  |  |  |
| T | 0 | FALSE | TRUE | TRUE | 25 |  | Positive | Negative | Quidel QuickVue |  |  |
| T | 1 |  | FALSE |  | 32 |  | Positive |  |  |  |  |
| T | 2 |  | FALSE |  | 33 |  | Positive |  |  |  |  |
| T | 3 |  | TRUE |  | 28 |  | Positive |  |  |  |  |
| U | -1 |  |  |  |  |  |  | Negative | Quidel QuickVue |  |  |
| U | 0 | FALSE | TRUE | TRUE | 25 |  | Positive | Negative | Quidel QuickVue |  |  |
| U | 1 |  | TRUE |  | 23 |  | Positive |  |  | Positive | Lucira Check-It |
| V | 0 | FALSE | TRUE | TRUE | 25 |  | Positive | Negative | Quidel QuickVue |  |  |
| W | 0 | FALSE | TRUE | TRUE | 25 |  | Positive | Negative | Abbott BinaxNOW | Positive | BioReference Laboratory |
| X* | 0 | FALSE | TRUE | TRUE | 28 |  | Positive | Negative | Quidel QuickVue |  |  |
| Y | 0 | FALSE | TRUE | TRUE | 28 |  | Positive | Negative | Quidel QuickVue |  |  |
| Y | 1 |  | FALSE |  | 32 |  | Positive |  |  |  |  |
| Z | 0 | FALSE | TRUE | TRUE | 28 |  | Positive | Negative | Quidel QuickVue |  |  |
| AA | 0 | FALSE | FALSE | FALSE | 30 |  | Positive | Negative | Quidel QuickVue |  |  |
| BB | 0 |  | FALSE |  | 35 |  | Positive |  |  |  |  |
| BB | 2 | FALSE | FALSE | FALSE | 33 |  | Positive | Negative | Quidel QuickVue |  |  |
| BB | 2 | FALSE | FALSE | FALSE | 30 |  | Positive | Negative | Quidel QuickVue |  |  |
| CC | -1 |  | TRUE |  |  |  |  | Negative |  |  |  |
| CC | 0 | FALSE | FALSE | FALSE | 34 |  | Positive | Negative |  |  |  |
| DD | -2 |  | FALSE |  |  |  | Negative |  |  |  |  |
| DD | -1 |  | FALSE |  |  |  |  |  |  | Negative | Lucira Check-It |
| DD | 0 | FALSE | TRUE | TRUE |  |  | Positive | Negative | Abbott BinaxNOW | Positive | Cue COVID-19 |

Notes: Cases are ordered from lowest to highest peak Ct among discordant paired tests. \*Individual with confirmed transmissions during the period of false negative antigen testing. \*\*High risk flag indicates an individual day when a person was observed to have infectious viral load and/or confirmed transmission event while testing negative on rapid antigen test. The index date (Day 0) was defined as the date of specimen collection for the first positive PCR test for an individual. The index date for Case S was based on symptom onset one day prior to first positive PCR. Dark shading indicates a “high risk” discordant matched pair of results having a negative antigen test during infectious viral load (saliva N-gene Ct <29); pale shading indicates a discordant matched pair of results with a negative antigen test and a positive PCR with viral load considered not to be infectious. PCR was the ThermoFisher Combo Kit. Some individuals additionally had nasal swabs collected for PCR testing and for those individuals the nasal Ct is also available to contextualize the saliva Ct.

